# Mental Health Impact of the First Wave of COVID-19 Pandemic on Spanish Healthcare Workers: a Large Cross-sectional Survey

**DOI:** 10.1101/2020.10.27.20220731

**Authors:** Jordi Alonso, Gemma Vilagut, Philippe Mortier, Montse Ferrer, Itxaso Alayo, Andrés Aragón-Peña, Enric Aragonès, Mireia Campos, Isabel del Cura-González, José I. Emparanza, Meritxell Espuga, M. Joao Forjaz, Ana González Pinto, Josep M. Haro, Nieves López Fresneña, Alma Martínez de Salázar, Juan D. Molina, Rafael M. Ortí Lucas, Mara Parellada, José Maria Pelayo-Terán, Aurora Pérez Zapata, José I. Pijoan, Nieves Plana, Teresa Puig, Cristina Rius, Carmen Rodriguez-Blazquez, Ferran Sanz, Consol Serra, Ronald C. Kessler, Ronny Bruffaerts, Eduard Vieta, Víctor Pérez-Solá, MINDCOVID Working group

**Author notes:** Corresponding author: Jordi Alonso, IMIM, CIBERESP, UPF. Address: IMIM. PRBB Building. Carrer del Doctor Aiguader 88. 08003 Barcelona, Spain. **The MINDCOVID Working Group** is formed by: Jordi Alonso, Itxaso Alayo, Manuel Alonso, Mar Álvarez, Benedikt Amann, Franco F. Amigo, Gerard Anmella, Andres Aragón, Nuria Aragonés, Enric Aragonès, Ana Isabel Arizón, Angel Asunsolo, Alfons Ayora, Laura Ballester, Puri Barbas, Josep Basora, Elena Bereciartua, Inés Bravo, Ignasi Bolibar, Xavier Bonfill, Alberto Cotillas, Andres Cuartero, Concha de Paz, Isabel del Cura, Maria Jesus del Yerro, Domingo Díaz, José Luis Domingo, Jose I. Emparanza, Mireia Espallargues, Meritxell Espuga, Patricia Estevan, M. Isabel Fernandez, Tania Fernandez, Montse Ferrer, Yolanda Ferreres, Giovanna Fico, M. Joao Forjaz, Rosa García-Barranco, J. Manuel García-Torrecillas C. García-Ribera, Araceli Garrido, Elisa Gil, Marta Gómez, Javier Gómez, Ana González-Pinto, Josep Maria Haro, Margarita Hernando, María Giola-Insigna, Milagros Iriberri, Nuria Jiménez, Xavi Jiménez, Amparo Larrauri, Fernando León, Nieves Lopez-Fresneña, Carmen López, Mayte López-Atanes Juan Antonio López-Rodríguez, Germán López-Cortacans, Alba Marcos, Jesús Martín, Vicente Martín, Mercedes Martínez-Cortés, Raquel Martínez-Martínez, Alma D. Martínez de Salazar, Isabel Martínez, Marco Marzola, Nelva Mata, Josep Maria Molina, Juan D. Molina, Emilia Molinero, Philippe Mortier, Carmen Muñoz, Andrea Murru, Jorge Olmedo, Rafael M Ortí, Rafael Padrós, Meritxell Pallejà, Raul Parra, Julio Pascual, José María Pelayo, Rosa Pla, Nieves Plana, Coro Pérez Aznar, Beatriz Pérez-Gómez, Aurora Pérez-Zapata, José Ignacio Pijoan, Elena Polentinos, Beatriz Puértolas, Maria Teresa Puig, Álex Quílez, M. Jesús Quintana, Antonio Quiroga, David Rentero, Cristina Rey, Cristina Rius, Carmen Rodríguez-Blázquez, M. José Rojas, Yamina Romero, Gabriel Rubio, Mercedes Rumayor, Pedro Ruiz, Margarita Sáenz, Jesus Sánchez, Ignacio Sánchez-Arcilla, Ferran Sanz, Consol Serra, Victoria Serra-Sutton, Manuela Serrano, Silvia Sola, Sara Solera, Miguel Soto, Alejandra Tarragoó Natividad Tolosa, Mireia Vázquez, Margarita Viciola, Eduard Vieta, Gemma Vilagut, Sara Yago, Jesus Yañez, Yolanda Zapico, Luis Maria Zorita, Iñaki Zorrilla, Saioa L. Zurbano, and Víctor Perez-Solá. **CONTRIBUTORS** JA, GV, and PM reviewed the literature. JA, GV, PM, MF, EA, VPS, JMH, RCK, and RB conceived and designed the study. EA, JDM, NL, TP, JMPT, JIP, JIE, ME, NP, AGP, CR, EA, ICG, AAP, MC, APZ, EV, CS, and VPS acquired the data. GV, IA, and PM cleaned and analyzed the data. JA, GV, and PM drafted the initial version of the manuscript. All authors reviewed the initial draft and made critical contribution to the interpretation of the data and approved the manuscript. The corresponding authors attest that all listed authors meet authorship criteria and that no others meeting the criteria have been omitted.

## Abstract

**Introduction:** Healthcare workers are vulnerable to adverse mental health impacts of COVID-19. We assessed prevalence of mental disorders and associated factors during the first wave of the pandemic among healthcare professionals in Spain.

**Methods:** All workers in 18 healthcare institutions (6 AACC) in Spain were invited to a series of online surveys assessing a wide range of individual characteristics, COVID-19 infection status and exposure, and mental health status. Here we report: current mental disorders (Major Depressive Disorder-MDD- [PHQ-8≥10], Generalized Anxiety Disorder-GAD- [GAD-7≥10], Panic attacks, Posttraumatic Stress Disorder –PTSD- [PCL-5≥7]; and Substance Use Disorder –SUD-[CAGE-AID≥2]. Severe disability assessed by the Sheehan Disability Scale was used to identify “*disabling*” current mental disorders.

**Results:** 9,138 healthcare workers participated. Prevalence of screen-positive disorder: 28.1% MDD; 22.5% GAD, 24.0% Panic; 22.2% PTSD; and 6.2% SUD. Overall 45.7% presented any current and 14.5% any disabling current mental disorder. Healthcare workers with prior lifetime mental disorders had almost twice the prevalence of current disorders than those without. Adjusting for all other variables, odds of any disabling mental disorder were: prior lifetime disorders (TUS: OR=5.74; 95%CI 2.53-13.03; Mood: OR=3.23; 95%CI:2.27-4.60; Anxiety: OR=3.03; 95%CI:2.53-3.62); age category 18-29 years (OR=1.36; 95%CI:1.02-1.82), caring “all of the time” for COVID-19 patients (OR=5.19; 95%CI: 3.61-7.46), female gender (OR=1.58; 95%CI: 1.27-1.96) and having being in quarantine or isolated (OR= 1.60; 95CI:1.31-1.95).

**Conclusions:** Current mental disorders were very frequent among Spanish healthcare workers during the first wave of COVID-19. As the pandemic enters its second wave, careful monitoring and support is needed for healthcare workers, especially those with previous mental disorders and those caring COVID-19 very often.

## INTRODUCTION

COVID-19 represents a major health challenge worldwide and several populations may experience adverse mental health related to the COVID-19 pandemic^1,2^. Among them, front-line healthcare workers are considered an extremely at risk population because of their direct exposure to infected patients, the limited availability of protective equipment, and the increased workload related to the pandemic. Compared to the general community, healthcare workers have about 12 times more risk for a positive COVID-19 test^3^. Although with noticeable regional and international variations, it is estimated that 10-20% of all COVID-19 diagnoses occur in this population segment^4,5^. In addition to the risk of contagion and insufficiency of equipment and health services preparedness there is great concern for the potential impact (acute and longer term) on the mental health of healthcare workers.

Several systematic reviews and meta-analyses including studies on health care workers have documented that the first wave of the COVID-19 was associated with an increase of symptoms of depression, anxiety, insomnia, and burnout, as well as other adverse psychosocial outcomes. Luo et al^6^, estimated that a quarter of healthcare workers suffered from anxiety (26%), depression (25%), and that about a third suffered substantial stress. Similar figures were reported in other systematic reviews^7-9^. In Spain, a number of studies have been carried out to assess mental health of healthcare workers during the first wave of the COVID-19 pandemic^10-14^. In general, results are consistent with international data, showing high levels of anxiety, depression and stress symptoms.

However, differences in study design, sample size as well as variation in the assessment and reporting of psychological impact and mental disorders hamper comparisons across studies. Importantly, current studies have limited value when it comes to assessing the needs for care associated with the impact of COVID-19 among healthcare workers. There is a necessity of credible and actionable indicators of mental disorders and their impact which more directly enable policy makers to allocate adequate resources when planning interventions.

Here we aimed to estimate the prevalence of clinically significant mental disorders among Spanish healthcare professionals during the first wave of the COVID-19 pandemic using a representative sample and well-validated screeners of common mental disorders. Specifically, our objectives were to estimate 1) prevalence of specific mental disorders, any such disorder, and any disabling disorder both in the total sample healthcare professionals and in subsamples of those with/without prior lifetime mental disorders; and 2) associations of individual and professional characteristics, COVID-19 infection status, and COVID-19 exposure with these mental disorders.

## METHODS

### Study design, population and sampling

A multicenter, prospective, observational cohort study of Spanish healthcare workers was carried out in a convenience sample of 18 health care institutions from 6 Autonomous Communities in Spain (i.e., Andalusia, the Basque Country, Castile and Leon, Catalonia, Madrid, and Valencia). Institutions were selected to reflect the geographical and sociodemographic variability in Spain; all participating centers came from regions with high COVID-19 caseloads. Here we report on the baseline assessment of the cohort, which consists of de-identified web-based self-report surveys administered soon after the first COVID-19 outbreak in Spain (May 5 – September 7, 2020).

In each participating health care institution, institutional representatives invited all employed hospital workers to participate using the hospitals’ administrative email distribution lists (i.e., census sampling). No further advertising of the survey was done and no incentives were offered for participation. The invitation email included an anonymous link to access the web-based survey platform (qualtrics.com). Median survey response time was 21.4 minutes (IQR 16.5-30.0).

Informed consent was obtained from all participants at the first survey page. Up to two reminder emails were sent within a 2-4 weeks period after the initial invitation. At the end of the survey, all participants were provided with a detailed list of local mental healthcare resources, including coordinates to nearby emergency care for respondents with a 30-day suicide attempt.

Participation was anonymous but participants could provide their email address at the end of the survey to participate in follow-up assessments of the cohort, which are conducted both at pre-specified time points, and in function of the course of the pandemic.

### Measures

#### Current Mental Disorders

– *Major Depressive Disorder (MDD):* evaluated with the Patient Health Questionnaire (PHQ-8). We used the Spanish version of the PHQ-8 (https://www.phqscreeners.com) with the cut-off point of 10 or higher of the sum score to indicate current MDD. The PHQ-8 shows high reliability (>0.8) and good diagnostic accuracy for Major Depressive Disorder (AUC>0.90)^15^.
– *Generalized Anxiety Disorder (GAD):* evaluated with the seven-item Generalized Anxiety Disorder scale (GAD-7), which has a good performance to detect anxiety (AUC>0.8^16^). We used the Spanish version of the GAD-7 (https://www.phqscreeners.com) and considered the cut-off point of 10 or higher to indicate a current GAD.
– *Panic attacks:* the number of panic attacks in the 30 days prior to the interview was assessed with an item from the World Mental Health-International College Student-WMH-ICS^17^. A dichotomous variable was created to indicate the presence of panic attacks.
– *Posttraumatic Stress Disorder (PTSD):* assessed using the 4-item version of the PTSD checklist for DSM-5 (PCL-5)^18-19^ which generates diagnoses that closely parallel those of the full PCL□5 (AUC>0.9), making it well□suited for screening^19^. We used the Spanish version of the questionnaire^20^, and considered a cut-off point of 7 to indicate current PTSD.
– *Substance Use Disorder (SUD):* evaluated with the CAGE-AID questionnaire, that consists of 4 items focusing on Cutting down, Annoyance by criticism, Guilty feeling, and Eye-openers. The CAGE-AID has been proved useful in helping to make a diagnosis of alcoholism^21^ and Substance Use Disorder^22^. The questionnaire has been adapted into Spanish. A cut-off point of 2 was considered to indicate current SUD^23^.
– *Disabling mental disorder*: a mental disorder was considered “disabling” if the participant reported severe role impairment during the past 12 months according to an adapted version of the Sheehan Disability Scale^24,25^. A 0–10 visual analogue scale was used to rate the degree of impairment for four domains: home management/chores, work, close personal relationships, and social life. The scale was labeled as no interference (0), mild (1–3), moderate (4–6), severe (7–9), and very severe (10) interference. Severe role impairment was defined as having a 7–10 rating^26-28^.
– *Prior lifetime mental disorders*: lifetime mental disorders prior to the onset of the COVID-19 outbreak were assessed using single-item screener variables based on the Composite International Diagnostic Interview (CIDI), including mood (i.e., depressive and bipolar disorders), anxiety (i.e., panic attacks, generalized anxiety and obsessive-compulsive disorders), substance use (i.e., alcohol, illicit drugs, and prescription drugs with or without prescription), and other disorders^28^.

#### COVID-19 exposure and infection status

We assessed the frequency of direct exposure to COVID-19 infected patients during professional activity using one 5-level Likert type item, ranging from “*none of the time*” to “*all of the time*. We defined *frontline healthcare workers* those reporting being exposed “*all of the time*” or “*most of the time*” to COVID-19 patients. We assessed COVID-19 infection status asking whether the respondent had been hospitalized for COVID-19 infection and/or had a positive COVID-19 test or medical diagnosis not requiring hospitalization. We also asked whether the respondent had been in isolation or quarantine because of exposure to COVID-19 infected person(s), and whether s/he had close ones infected with COVID-19.

#### Individual characteristics

We assessed: age; gender; country of birth; marital status; having children in care; living situation; and profession into 5 categories: medical doctors, nurses, auxiliary nurses, other professions involved in patient care, and other professions not involved in patient care.

### Ethical considerations

The study complies with the principles established by national and international regulations, including the Declaration of Helsinki and the Code of Ethics. The data were pseudo-anonymized through encrypted identifiers, separating the personal information from the rest of the study data, to guarantee privacy and ensure de-identified treatment of the data in the analysis. The study protocol was approved by the IRB Parc de Salut Mar (2020/9203/I) and by the corresponding IRBs of all the participating centers. The study is registered at ClinicalTrials.gov (https://clinicaltrials.gov/ct2/show/NCT04556565).

### Statistical analysis

Analyses were restricted to the n = 9,146 respondents who completed all Mental Health items of the questionnaire. Of them, an additional n = 8 respondents were not included because they did not identify themselves with neither male nor female gender. In order to improve representativeness, observed data were weighted using raking procedure to reproduce marginal distributions of gender, age and professional category of healthcare personnel in each participating institution, as well as distribution of personnel across institutions.

Median proportion of missingness per variable was less than 1%. However, to optimize survey timings, the Sheehan disability scale was assessed only in a random 60% of the sample. Missing item-level data from the Sheehan scale and from all other variables included in the analysis were handled using multiple imputation (MI) by chained equations with 40 imputed datasets and 10 iterations. These included all study variables and additional variables from the questionnaire as predictors in the imputation regression equations. Pooled estimates from multiple imputations and MI-based standard errors taking into account within-imputation and between-imputation variances were obtained.

Distribution of individual characteristics and COVID-19 infection and exposure variables were obtained for the whole sample as weighted percentage and standard error. Prevalence estimates of specific current mental disorders, any current mental disorder, and any disabling disorder were estimated, overall and stratified by individual characteristics. Chi-square tests from MI pooled using Rubin’s rule were used to determine significant differences across strata. Adjustment for multiple comparisons was performed using the Benjamini–Hochberg procedure^29^ with a false discovery rate of 5%. Bivariable associations between each individual characteristic and current mental disorders and severe mental disorder were estimated for the overall sample, and separately for individuals without a history of prior lifetime mental disorders (*new onset)*, and for those reporting prior mental disorders *(persistence/relapse)*. Odds ratios (OR) and MI-based 95% confidence intervals (CIs) for each characteristic were calculated with logistic regression, adjusted by week of survey and health center membership. Finally, multivariable associations between all COVID-19 exposure and infection status, individual characteristics considered and current and disabling mental disorders were also estimated with logistic regression, stratifying by prior lifetime mental disorders.

MI were carried out using package mice from R^30,31^. Analyses were performed using R v3.4.2^32^ and SAS v9.4^33^.

## RESULTS

### Survey response

A total of 9,138 healthcare workers participated in the surveys. The *response rate* is difficult to estimate given that the survey *view rate* (i.e., the proportion of hospital workers that opened the invitation email) is unknown, except for one hospital (26.4%). The *survey participation rate* (i.e., those that agreed to participate divided by those that responded to the informed consent on the first survey page) was 89.0%, and the *survey completion rate* (i.e., those that completed the survey among those that agreed to participate) was 80.8%. When the denominator used to calculate the response rate is the total number of health care workers listed in the email distribution list or the total number of healthcare workers employed as provided by the hospital representatives, the survey response is 12.5% (see **supplementary Table 1**).

### Prevalence of current mental disorders

The first two columns of **Table 1** show the size and weighted distribution of the sample studied. Healthcare professionals were mostly female (77.3%), the larger age group was 30-49 years (45.8%), just over half were married (53.0%), four out of ten were living with children (41.4%), and 57.2% were living in an apartment. About a fourth (26.4%) were physicians, and 30.6% were nurses, and the majority were working in a hospital (54.1%). Almost 80% of participants were directly involved in patient care, although less than a half (43.6%) were directly exposed to COVID-19 patients *all* or *most of the time* (i. e., frontline workers). Almost a fifth (17.4%) had COVID-19, 13.8% had their spouse/couple, children or parents infected with COVID-19, and up to 25.5% had been isolated or quarantined. An important proportion (41.6%) reported a lifetime mental disorders before the pandemic.

**Table 1.**
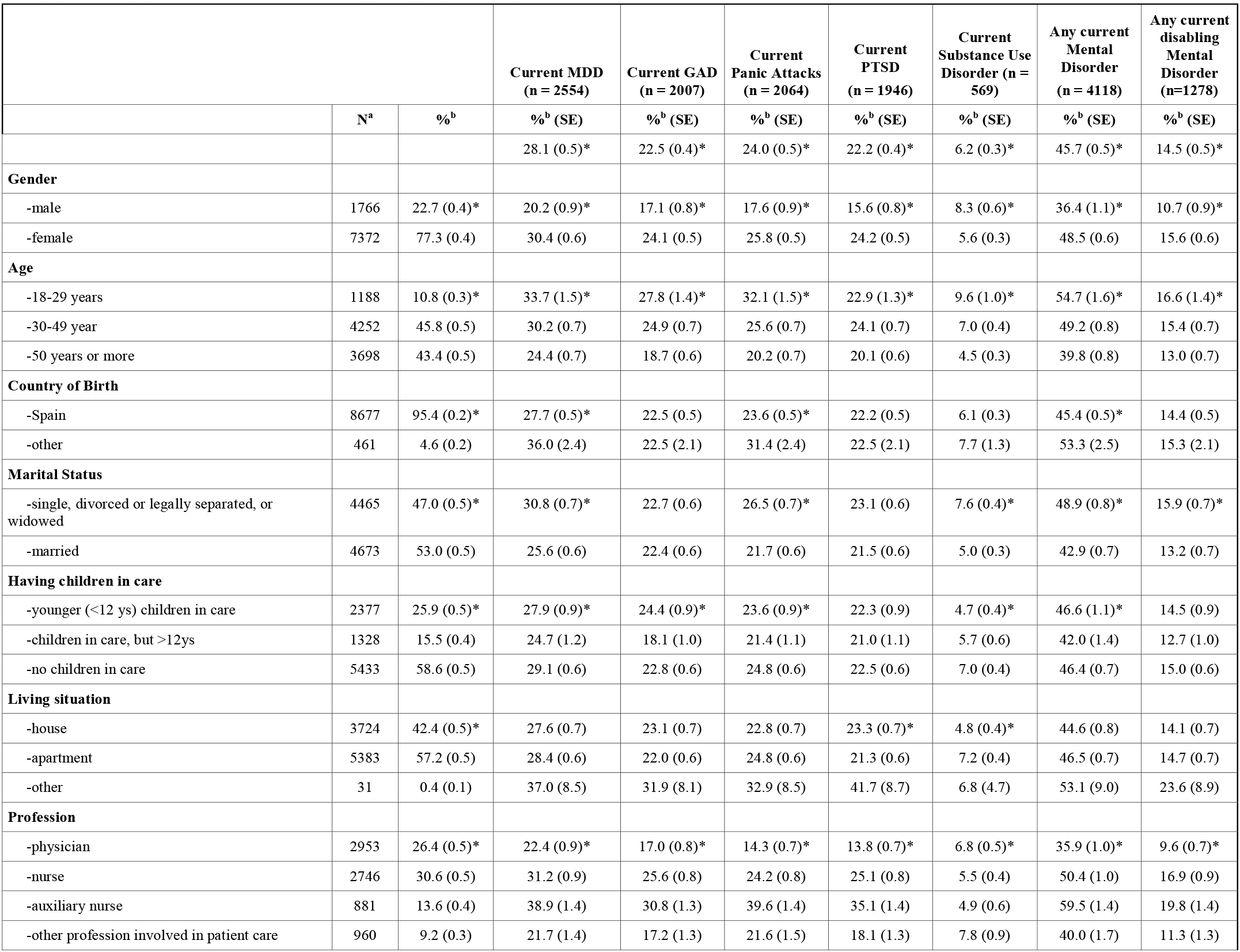
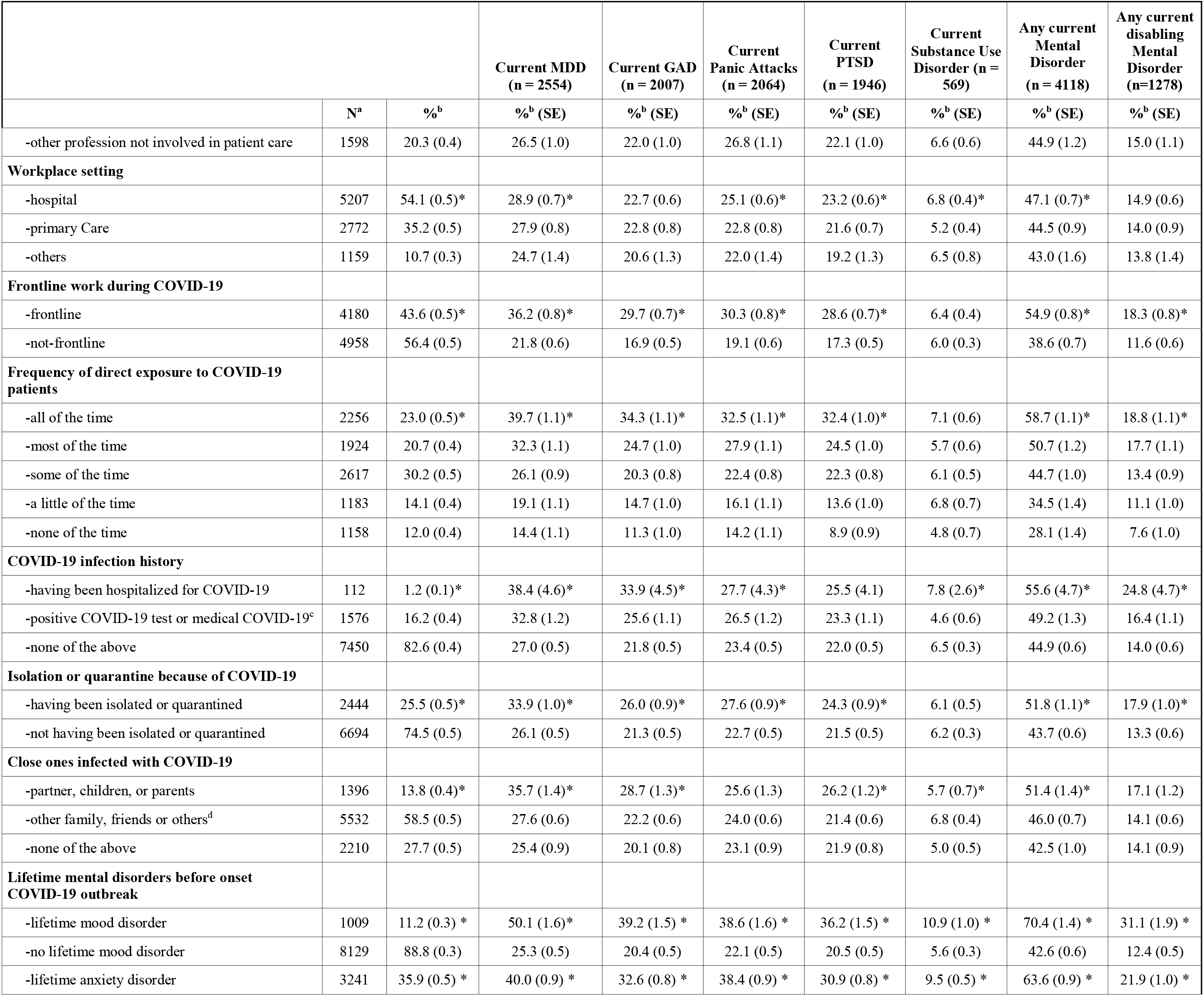
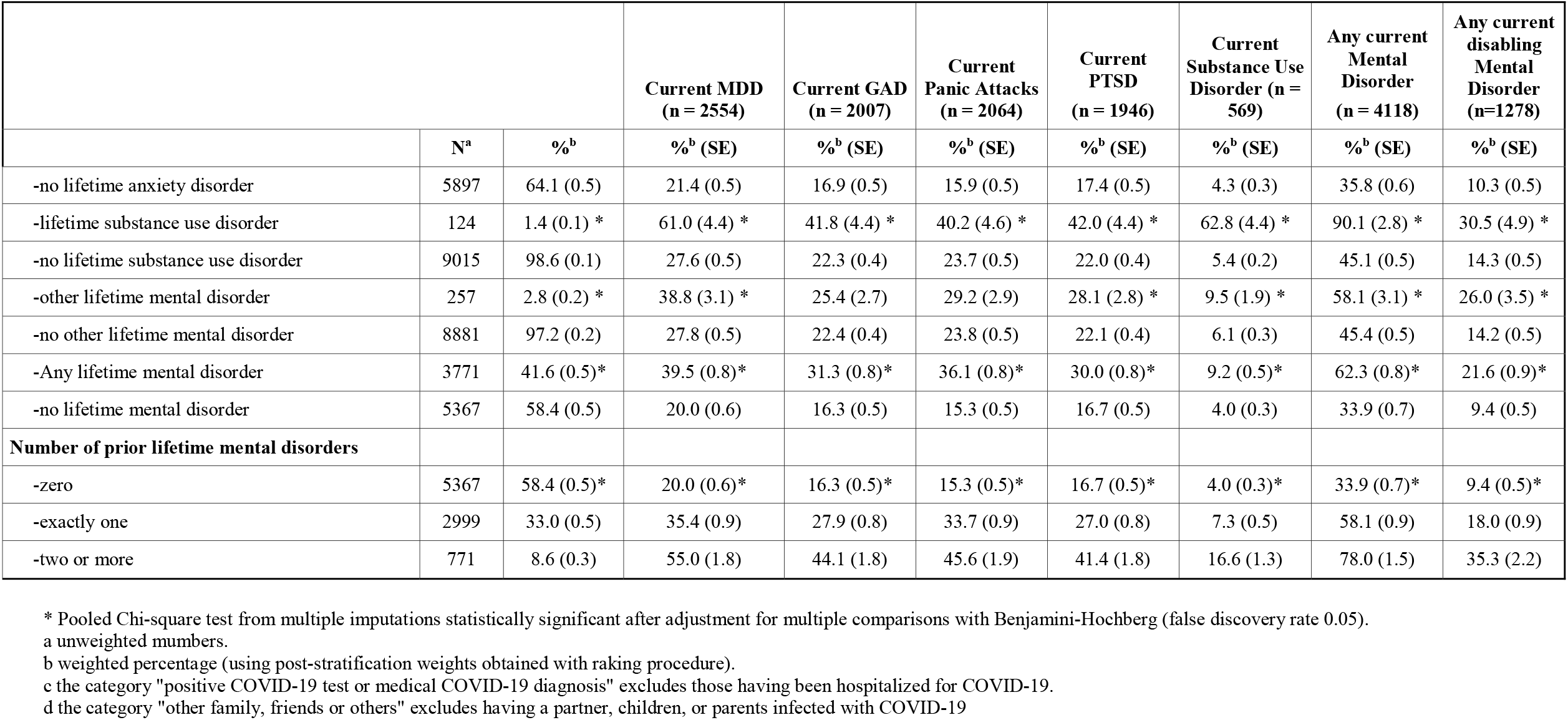
Prevalence of current mental disorders among Spanish healthcare workers, according to individual characteristics, COVID-19 exposure, and prior lifetime disorders. MINCOVID study (N =9,138) (absolute numbers and weighted proportions).

|  |  |  | **Current MDD**<br>**(n = 2554)** | **Current GAD**<br>**(n = 2007)** | **Current**<br>**Panic Attacks**<br>**(n = 2064)** | **Current**<br>**PTSD**<br>**(n = 1946)** | **Current**<br>**Substance Use**<br>**Disorder (n =**<br>**569)** | **Any current**<br>**Mental**<br>**Disorder**<br>**(n = 4118)** | **Any current**<br>**disabling**<br>**Mental**<br>**Disorder**<br>**(n=1278)** |
| --- | --- | --- | --- | --- | --- | --- | --- | --- | --- |
|  | **N<sup>a</sup>** | **%<sup>b</sup>** | **%<sup>b</sup> (SE)** | **%<sup>b</sup> (SE)** | **%<sup>b</sup> (SE)** | **%<sup>b</sup> (SE)** | **%<sup>b</sup> (SE)** | **%<sup>b</sup> (SE)** | **%<sup>b</sup> (SE)** |
|  |  |  | 28.1 (0.5)* | 22.5 (0.4)* | 24.0 (0.5)* | 22.2 (0.4)* | 6.2 (0.3)* | 45.7 (0.5)* | 14.5 (0.5)* |
| **Gender** |  |  |  |  |  |  |  |  |  |
| -male | 1766 | 22.7 (0.4)* | 20.2 (0.9)* | 17.1 (0.8)* | 17.6 (0.9)* | 15.6 (0.8)* | 8.3 (0.6)* | 36.4 (1.1)* | 10.7 (0.9)* |
| -female | 7372 | 77.3 (0.4) | 30.4 (0.6) | 24.1 (0.5) | 25.8 (0.5) | 24.2 (0.5) | 5.6 (0.3) | 48.5 (0.6) | 15.6 (0.6) |
| **Age** |  |  |  |  |  |  |  |  |  |
| -18-29 years | 1188 | 10.8 (0.3)* | 33.7 (1.5)* | 27.8 (1.4)* | 32.1 (1.5)* | 22.9 (1.3)* | 9.6 (1.0)* | 54.7 (1.6)* | 16.6 (1.4)* |
| -30-49 year | 4252 | 45.8 (0.5) | 30.2 (0.7) | 24.9 (0.7) | 25.6 (0.7) | 24.1 (0.7) | 7.0 (0.4) | 49.2 (0.8) | 15.4 (0.7) |
| -50 years or more | 3698 | 43.4 (0.5) | 24.4 (0.7) | 18.7 (0.6) | 20.2 (0.7) | 20.1 (0.6) | 4.5 (0.3) | 39.8 (0.8) | 13.0 (0.7) |
| **Country of Birth** |  |  |  |  |  |  |  |  |  |
| -Spain | 8677 | 95.4 (0.2)* | 27.7 (0.5)* | 22.5 (0.5) | 23.6 (0.5)* | 22.2 (0.5) | 6.1 (0.3) | 45.4 (0.5)* | 14.4 (0.5) |
| -other | 461 | 4.6 (0.2) | 36.0 (2.4) | 22.5 (2.1) | 31.4 (2.4) | 22.5 (2.1) | 7.7 (1.3) | 53.3 (2.5) | 15.3 (2.1) |
| **Marital Status** |  |  |  |  |  |  |  |  |  |
| -single, divorced or legally separated, or<br>widowed | 4465 | 47.0 (0.5)* | 30.8 (0.7)* | 22.7 (0.6) | 26.5 (0.7)* | 23.1 (0.6) | 7.6 (0.4)* | 48.9 (0.8)* | 15.9 (0.7)* |
| -married | 4673 | 53.0 (0.5) | 25.6 (0.6) | 22.4 (0.6) | 21.7 (0.6) | 21.5 (0.6) | 5.0 (0.3) | 42.9 (0.7) | 13.2 (0.7) |
| **Having children in care** |  |  |  |  |  |  |  |  |  |
| -younger (<12 ys) children in care | 2377 | 25.9 (0.5)* | 27.9 (0.9)* | 24.4 (0.9)* | 23.6 (0.9)* | 22.3 (0.9) | 4.7 (0.4)* | 46.6 (1.1)* | 14.5 (0.9) |
| -children in care, but >12ys | 1328 | 15.5 (0.4) | 24.7 (1.2) | 18.1 (1.0) | 21.4 (1.1) | 21.0 (1.1) | 5.7 (0.6) | 42.0 (1.4) | 12.7 (1.0) |
| -no children in care | 5433 | 58.6 (0.5) | 29.1 (0.6) | 22.8 (0.6) | 24.8 (0.6) | 22.5 (0.6) | 7.0 (0.4) | 46.4 (0.7) | 15.0 (0.6) |
| **Living situation** |  |  |  |  |  |  |  |  |  |
| -house | 3724 | 42.4 (0.5)* | 27.6 (0.7) | 23.1 (0.7) | 22.8 (0.7) | 23.3 (0.7)* | 4.8 (0.4)* | 44.6 (0.8) | 14.1 (0.7) |
| -apartment | 5383 | 57.2 (0.5) | 28.4 (0.6) | 22.0 (0.6) | 24.8 (0.6) | 21.3 (0.6) | 7.2 (0.4) | 46.5 (0.7) | 14.7 (0.7) |
| -other | 31 | 0.4 (0.1) | 37.0 (8.5) | 31.9 (8.1) | 32.9 (8.5) | 41.7 (8.7) | 6.8 (4.7) | 53.1 (9.0) | 23.6 (8.9) |
| **Profession** |  |  |  |  |  |  |  |  |  |
| -physician | 2953 | 26.4 (0.5)* | 22.4 (0.9)* | 17.0 (0.8)* | 14.3 (0.7)* | 13.8 (0.7)* | 6.8 (0.5)* | 35.9 (1.0)* | 9.6 (0.7)* |
| -nurse | 2746 | 30.6 (0.5) | 31.2 (0.9) | 25.6 (0.8) | 24.2 (0.8) | 25.1 (0.8) | 5.5 (0.4) | 50.4 (1.0) | 16.9 (0.9) |
| -auxiliary nurse | 881 | 13.6 (0.4) | 38.9 (1.4) | 30.8 (1.3) | 39.6 (1.4) | 35.1 (1.4) | 4.9 (0.6) | 59.5 (1.4) | 19.8 (1.4) |
| -other profession involved in patient care | 960 | 9.2 (0.3) | 21.7 (1.4) | 17.2 (1.3) | 21.6 (1.5) | 18.1 (1.3) | 7.8 (0.9) | 40.0 (1.7) | 11.3 (1.3) |

|  |  |  | Current MDD<br>(n = 2554) | Current GAD<br>(n = 2007) | Current<br>Panic Attacks<br>(n = 2064) | Current<br>PTSD<br>(n = 1946) | Current<br>Substance Use<br>Disorder (n =<br>569) | Any current<br>Mental<br>Disorder<br>(n = 4118) | Any current<br>disabling<br>Mental<br>Disorder<br>(n=1278) |
| --- | --- | --- | --- | --- | --- | --- | --- | --- | --- |
|  | N<sup>a</sup> | %<sup>b</sup> | %<sup>b</sup> (SE) | %<sup>b</sup> (SE) | %<sup>b</sup> (SE) | %<sup>b</sup> (SE) | %<sup>b</sup> (SE) | %<sup>b</sup> (SE) | %<sup>b</sup> (SE) |
| -other profession not involved in patient care | 1598 | 20.3 (0.4) | 26.5 (1.0) | 22.0 (1.0) | 26.8 (1.1) | 22.1 (1.0) | 6.6 (0.6) | 44.9 (1.2) | 15.0 (1.1) |
| **Workplace setting** |  |  |  |  |  |  |  |  |  |
| -hospital | 5207 | 54.1 (0.5)* | 28.9 (0.7)* | 22.7 (0.6) | 25.1 (0.6)* | 23.2 (0.6)* | 6.8 (0.4)* | 47.1 (0.7)* | 14.9 (0.6) |
| -primary Care | 2772 | 35.2 (0.5) | 27.9 (0.8) | 22.8 (0.8) | 22.8 (0.8) | 21.6 (0.7) | 5.2 (0.4) | 44.5 (0.9) | 14.0 (0.9) |
| -others | 1159 | 10.7 (0.3) | 24.7 (1.4) | 20.6 (1.3) | 22.0 (1.4) | 19.2 (1.3) | 6.5 (0.8) | 43.0 (1.6) | 13.8 (1.4) |
| **Frontline work during COVID-19** |  |  |  |  |  |  |  |  |  |
| -frontline | 4180 | 43.6 (0.5)* | 36.2 (0.8)* | 29.7 (0.7)* | 30.3 (0.8)* | 28.6 (0.7)* | 6.4 (0.4) | 54.9 (0.8)* | 18.3 (0.8)* |
| -not-frontline | 4958 | 56.4 (0.5) | 21.8 (0.6) | 16.9 (0.5) | 19.1 (0.6) | 17.3 (0.5) | 6.0 (0.3) | 38.6 (0.7) | 11.6 (0.6) |
| **Frequency of direct exposure to COVID-19 patients** |  |  |  |  |  |  |  |  |  |
| -all of the time | 2256 | 23.0 (0.5)* | 39.7 (1.1)* | 34.3 (1.1)* | 32.5 (1.1)* | 32.4 (1.0)* | 7.1 (0.6) | 58.7 (1.1)* | 18.8 (1.1)* |
| -most of the time | 1924 | 20.7 (0.4) | 32.3 (1.1) | 24.7 (1.0) | 27.9 (1.1) | 24.5 (1.0) | 5.7 (0.6) | 50.7 (1.2) | 17.7 (1.1) |
| -some of the time | 2617 | 30.2 (0.5) | 26.1 (0.9) | 20.3 (0.8) | 22.4 (0.8) | 22.3 (0.8) | 6.1 (0.5) | 44.7 (1.0) | 13.4 (0.9) |
| -a little of the time | 1183 | 14.1 (0.4) | 19.1 (1.1) | 14.7 (1.0) | 16.1 (1.1) | 13.6 (1.0) | 6.8 (0.7) | 34.5 (1.4) | 11.1 (1.0) |
| -none of the time | 1158 | 12.0 (0.4) | 14.4 (1.1) | 11.3 (1.0) | 14.2 (1.1) | 8.9 (0.9) | 4.8 (0.7) | 28.1 (1.4) | 7.6 (1.0) |
| **COVID-19 infection history** |  |  |  |  |  |  |  |  |  |
| -having been hospitalized for COVID-19 | 112 | 1.2 (0.1)* | 38.4 (4.6)* | 33.9 (4.5)* | 27.7 (4.3)* | 25.5 (4.1) | 7.8 (2.6)* | 55.6 (4.7)* | 24.8 (4.7)* |
| -positive COVID-19 test or medical COVID-19<sup>c</sup> | 1576 | 16.2 (0.4) | 32.8 (1.2) | 25.6 (1.1) | 26.5 (1.2) | 23.3 (1.1) | 4.6 (0.6) | 49.2 (1.3) | 16.4 (1.1) |
| -none of the above | 7450 | 82.6 (0.4) | 27.0 (0.5) | 21.8 (0.5) | 23.4 (0.5) | 22.0 (0.5) | 6.5 (0.3) | 44.9 (0.6) | 14.0 (0.6) |
| **Isolation or quarantine because of COVID-19** |  |  |  |  |  |  |  |  |  |
| -having been isolated or quarantined | 2444 | 25.5 (0.5)* | 33.9 (1.0)* | 26.0 (0.9)* | 27.6 (0.9)* | 24.3 (0.9)* | 6.1 (0.5) | 51.8 (1.1)* | 17.9 (1.0)* |
| -not having been isolated or quarantined | 6694 | 74.5 (0.5) | 26.1 (0.5) | 21.3 (0.5) | 22.7 (0.5) | 21.5 (0.5) | 6.2 (0.3) | 43.7 (0.6) | 13.3 (0.6) |
| **Close ones infected with COVID-19** |  |  |  |  |  |  |  |  |  |
| -partner, children, or parents | 1396 | 13.8 (0.4)* | 35.7 (1.4)* | 28.7 (1.3)* | 25.6 (1.3) | 26.2 (1.2)* | 5.7 (0.7)* | 51.4 (1.4)* | 17.1 (1.2) |
| -other family, friends or others<sup>d</sup> | 5532 | 58.5 (0.5) | 27.6 (0.6) | 22.2 (0.6) | 24.0 (0.6) | 21.4 (0.6) | 6.8 (0.4) | 46.0 (0.7) | 14.1 (0.6) |
| -none of the above | 2210 | 27.7 (0.5) | 25.4 (0.9) | 20.1 (0.8) | 23.1 (0.9) | 21.9 (0.8) | 5.0 (0.5) | 42.5 (1.0) | 14.1 (0.9) |
| **Lifetime mental disorders before onset COVID-19 outbreak** |  |  |  |  |  |  |  |  |  |
| -lifetime mood disorder | 1009 | 11.2 (0.3) * | 50.1 (1.6)* | 39.2 (1.5) * | 38.6 (1.6) * | 36.2 (1.5) * | 10.9 (1.0) * | 70.4 (1.4) * | 31.1 (1.9) * |
| -no lifetime mood disorder | 8129 | 88.8 (0.3) | 25.3 (0.5) | 20.4 (0.5) | 22.1 (0.5) | 20.5 (0.5) | 5.6 (0.3) | 42.6 (0.6) | 12.4 (0.5) |
| -lifetime anxiety disorder | 3241 | 35.9 (0.5) * | 40.0 (0.9) * | 32.6 (0.8) * | 38.4 (0.9) * | 30.9 (0.8) * | 9.5 (0.5) * | 63.6 (0.9) * | 21.9 (1.0) * |
|  | **N<sup>a</sup>** | **%<sup>b</sup>** | **%<sup>b</sup> (SE)** | **%<sup>b</sup> (SE)** | **%<sup>b</sup> (SE)** | **%<sup>b</sup> (SE)** | **%<sup>b</sup> (SE)** | **%<sup>b</sup> (SE)** | **%<sup>b</sup> (SE)** |
| -no lifetime anxiety disorder | 5897 | 64.1 (0.5) | 21.4 (0.5) | 16.9 (0.5) | 15.9 (0.5) | 17.4 (0.5) | 4.3 (0.3) | 35.8 (0.6) | 10.3 (0.5) |
| -lifetime substance use disorder | 124 | 1.4 (0.1) * | 61.0 (4.4) * | 41.8 (4.4) * | 40.2 (4.6) * | 42.0 (4.4) * | 62.8 (4.4) * | 90.1 (2.8) * | 30.5 (4.9) * |
| -no lifetime substance use disorder | 9015 | 98.6 (0.1) | 27.6 (0.5) | 22.3 (0.4) | 23.7 (0.5) | 22.0 (0.4) | 5.4 (0.2) | 45.1 (0.5) | 14.3 (0.5) |
| -other lifetime mental disorder | 257 | 2.8 (0.2) * | 38.8 (3.1) * | 25.4 (2.7) | 29.2 (2.9) | 28.1 (2.8) * | 9.5 (1.9) * | 58.1 (3.1) * | 26.0 (3.5) * |
| -no other lifetime mental disorder | 8881 | 97.2 (0.2) | 27.8 (0.5) | 22.4 (0.4) | 23.8 (0.5) | 22.1 (0.4) | 6.1 (0.3) | 45.4 (0.5) | 14.2 (0.5) |
| -Any lifetime mental disorder | 3771 | 41.6 (0.5)* | 39.5 (0.8)* | 31.3 (0.8)* | 36.1 (0.8)* | 30.0 (0.8)* | 9.2 (0.5)* | 62.3 (0.8)* | 21.6 (0.9)* |
| -no lifetime mental disorder | 5367 | 58.4 (0.5) | 20.0 (0.6) | 16.3 (0.5) | 15.3 (0.5) | 16.7 (0.5) | 4.0 (0.3) | 33.9 (0.7) | 9.4 (0.5) |
| **Number of prior lifetime mental disorders** |  |  |  |  |  |  |  |  |  |
| -zero | 5367 | 58.4 (0.5)* | 20.0 (0.6)* | 16.3 (0.5)* | 15.3 (0.5)* | 16.7 (0.5)* | 4.0 (0.3)* | 33.9 (0.7)* | 9.4 (0.5)* |
| -exactly one | 2999 | 33.0 (0.5) | 35.4 (0.9) | 27.9 (0.8) | 33.7 (0.9) | 27.0 (0.8) | 7.3 (0.5) | 58.1 (0.9) | 18.0 (0.9) |
| -two or more | 771 | 8.6 (0.3) | 55.0 (1.8) | 44.1 (1.8) | 45.6 (1.9) | 41.4 (1.8) | 16.6 (1.3) | 78.0 (1.5) | 35.3 (2.2) |

\* Pooled Chi-square test from multiple imputations statistically significant after adjustment for multiple comparisons with Benjamini-Hochberg (false discovery rate 0.05).
a unweighted numbers.
b weighted percentage (using post-stratification weights obtained with raking procedure).
c the category "positive COVID-19 test or medical COVID-19 diagnosis" excludes those having been hospitalized for COVID-19.
d the category "other family, friends or others" excludes having a partner, children, or parents infected with COVID-19

Also in **Table 1**, the prevalence of current mental disorders is presented according to the above variables. Overall, 28.1% met criteria for Major Depressive Disorder, between 22.2% and 24.0% met criteria for anxiety disorders (GAD, Panic attacks, or PTSD), and 6.2% met criteria for substance use disorder. In all, almost half of the sample (45.7%) met criteria for current mental disorder and about one in seven (14.5%) had a current disabling mental disorder.

The prevalence of any current mental disorder was significantly higher among healthcare workers with female gender, younger age, not born in Spain, not being married, or living with children less than 12 years of age or not having children at home. Auxiliary nurses and nurses showed the highest prevalence of current mental disorders (59.5% and 50.4%, respectively). There was a clear positive trend with higher exposure to COVID-19 patients, and those having the disease -- in particular those 112 professionals who had been hospitalized for COVID-19, having been isolated or quarantined, and whose parents, children or partner were infected with COVID-19. Prior lifetime mental disorders were strongly associated with presenting current mental disorder (especially those reporting previous substance use disorder or depression). The higher the number of prior lifetime mental disorders reported, the more likely the prevalence of any current disorder. Similar prevalence differences were found when considering current disabling mental disorders.

### Current mental disorders according to prior lifetime mental disorders

**Figure 1** shows current prevalence of mental disorders according to pre-COVID-19 pandemic lifetime mental disorders. Prevalence was consistently lower among workers without prior mental disorders (new onset), i.e., approximately half than among workers with prior mental disorders (persistent/relapsing).

**Figure 1.**
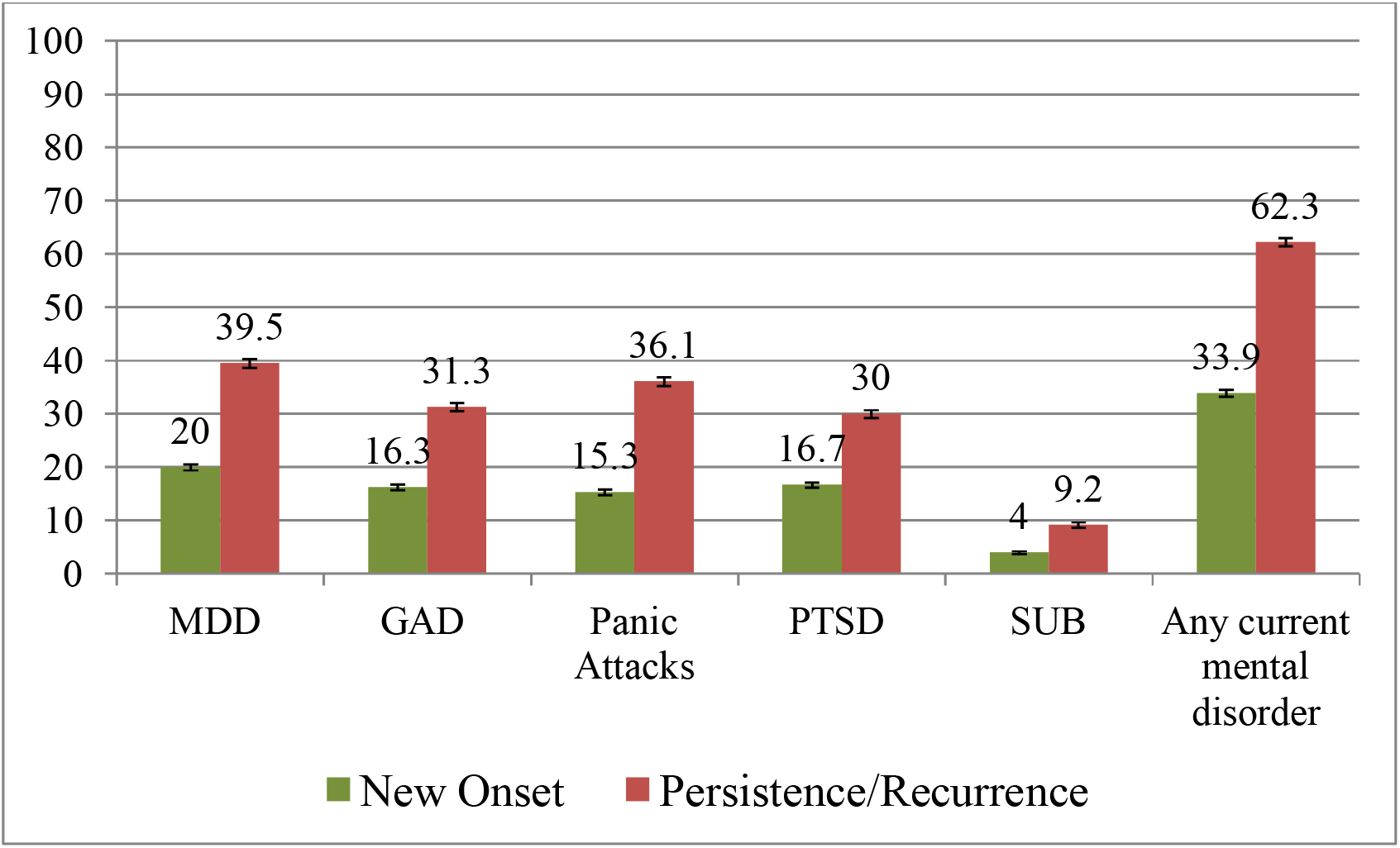
Current prevalence of mental disorders among Spanish healthcare workers during the first wave of the COVID-19 pandemic, according to prior lifetime mental disorders. MINDCOVID study (n =9,138). **Green bar**: workers with no pre-pandemic mental disorders (*new onset)*; **Red Bar**: workers with lifetime history of mental disorders (*persistence/recurrence*). MDD: Major Depressive Disorder; GAD: Generalized Anxiety Disorder; PTSD: Post-Stress Traumatic Disorder; SUB: Substance Use Disorder.

**Figure 2** shows current prevalence of any mental disorders (both disabling and non-disabling), according to pre-COVID-19 pandemic prior lifetime mental disorders. Among workers without prior mental disorders, the prevalence of any mental disorder (new onset) was almost 34% and one in four of those were disabling mental disorders (**Figure 2**.**A**). Among healthcare workers with any prior lifetime disorder, the prevalence of current disorders (persistence/relapse) was much higher (61%), and more frequently disabling (i.e., one in three) (**Figure 2**.**B**).

**Figure 2.**
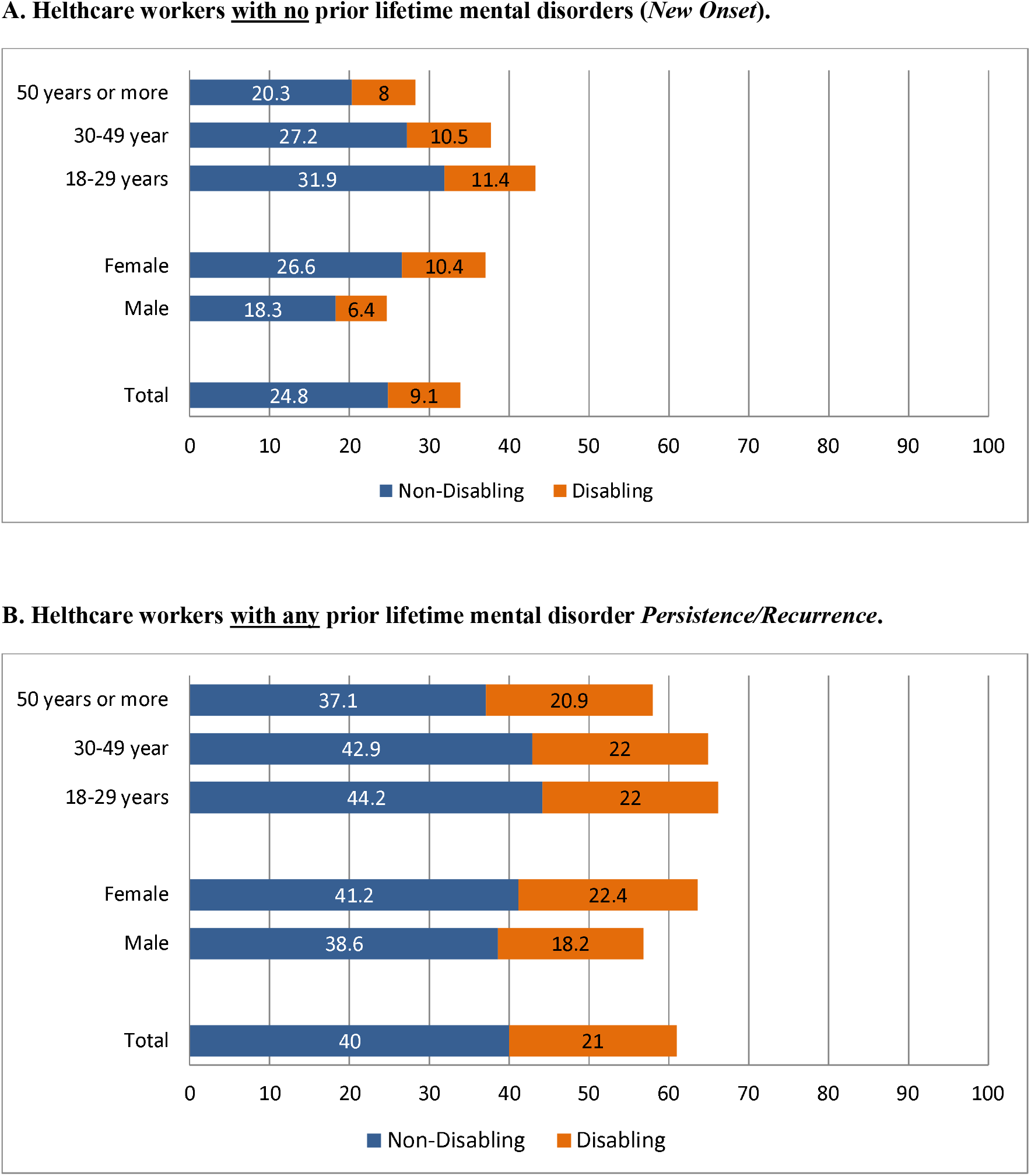
Current prevalence of any mental disorders (disabling and non-disabling) among Spanish healthcare workers during the first wave of the COVID-19 pandemic, according the pre-COVID-19 pandemic lifetime mental disorders and individual characteristics. MINDCOVID study (n =9,138).

### Factors associated with current mental disorders

**Table 2** shows bivariate associations of individual characteristics, personal COVID-19 exposure and prior lifetime mental disorders with any current mental disorder and with any current disabling mental disorder. The first two columns present the associations for the overall sample (n=9,138) that had been presented in table 1 in the form of Odds Ratios, once adjusting by week of the survey and by healthcare center. **Table 2** also shows these associations, stratifying by prior lifetime mental disorders. Columns 3-4 present data for those with no mental disorders prior to the COVID-19 pandemic and columns 5-6 refer to those reporting mental disorders before the first wave of the COVID-19 pandemic. In general, all the above-mentioned variables under study with any current (disabling) mental disorders were significantly associated with both new onset and persistence/relapse mental disorders. However, the association of hospitalization due to COVID-19 with any current disabling disorder was only significant for those with previous mental disorders. Among those with previous mental disorders, previous SUD and previous depression were most strongly associated with current persisting/relapsing mental disorders.

**Table 2.** Bivariate associations between individual characteristics, COVID-19 exposure, and prior lifetime (LT) mental disorders with any current and any disabling mental disorders. Spanish healthcare workers, MINDCOVID study (N=9,138).

|  | ALL (n=9,138) |  | No Prior LT Mental Disorders<br>(New Onset) (n=5,367) |  | Prior LT Mental Disorder<br>(Persistence Relapse) (n=3,771) |  |
| --- | --- | --- | --- | --- | --- | --- |
|  | Any current Mental Disorder (n=4,118) | Any current disabling Mental disorder (n=1,278) | Any current Mental Disorder (n=1,818) | Any current disabling Mental disorder (n=485) | Any current Mental Disorder (n=2,300) | Any current disabling mental disorder (n=793) |
|  | OR (95%CI) | OR (95%CI) | OR (95%CI) | OR (95%CI) | OR (95%CI) | OR (95%CI) |
| Gender - female (vs male) | 1.60 (1.44-1.78)* | 1.77 (1.46-2.14)* | 1.74 (1.50-2.02)* | 1.96 (1.43-2.70)* | 1.32 (1.12-1.56)* | 1.45 (1.11-1.88)* |
| Age |  |  |  |  |  |  |
| -18-29 years | 1.77 (1.53-2.05)* | 1.61 (1.27-2.03)* | 1.89 (1.54-2.34)* | 1.73 (1.17-2.56)* | 1.36 (1.09-1.69)* | 1.22 (0.89-1.67) |
| -30-49 year | 1.48 (1.35-1.62)* | 1.39 (1.19-1.63)* | 1.55 (1.37-1.76)* | 1.51 (1.19-1.91)* | 1.35 (1.17-1.57)* | 1.25 (1.00-1.57) |
| -50 years or more | Ref. | Ref. | Ref. | Ref. | Ref. | Ref. |
| Country of Birth -Spain (vs Other) | 0.74 (0.60-0.91)* | 0.85 (0.61-1.19) | 0.80 (0.60-1.06) | 0.93 (0.54-1.62) | 0.69 (0.49-0.97)* | 0.83 (0.51-1.35) |
| Marital status - married (vs single, divorced or legally separated, or widowed) | 0.81 (0.74-0.88)* | 0.77 (0.67-0.89)* | 0.88 (0.78-0.99)* | 0.88 (0.70-1.10) | 0.86 (0.75-0.99)* | 0.82 (0.67-1.00) |
| Having children in care |  |  |  |  |  |  |
| -younger (<12 ys) children in care | 1.04 (0.94-1.15) | 1.01 (0.86-1.18) | 1.17 (1.02-1.34)* | 1.18 (0.91-1.51) | 1.00 (0.85-1.18) | 0.96 (0.76-1.21) |
| -children in care, but >12ys | 0.86 (0.76-0.97)* | 0.81 (0.66-0.99)* | 0.89 (0.75-1.06) | 0.80 (0.58-1.10) | 0.90 (0.74-1.10) | 0.89 (0.66-1.18) |
| -no children in care | Ref. | Ref. | Ref. | Ref. | Ref. | Ref. |
| Living situation |  |  |  |  |  |  |
| -house | 0.78 (0.38-1.61) | 0.53 (0.19-1.43) | 0.74 (0.23-2.40) | 0.45 (0.09-2.29) | 1.16 (0.45-3.00) | 0.79 (0.21-3.00) |
| -apartment | 0.85 (0.41-1.75) | 0.56 (0.20-1.53) | 0.80 (0.25-2.57) | 0.47 (0.09-2.40) | 1.17 (0.45-3.02) | 0.78 (0.20-3.01) |
| -other | Ref. | Ref. | Ref. | Ref. | Ref. | Ref. |
| Profession |  |  |  |  |  |  |
| -physician | 0.63 (0.55-0.71)* | 0.50 (0.40-0.62)* | 0.66 (0.55-0.79)* | 0.47 (0.33-0.68)* | 0.64 (0.52-0.78)* | 0.53 (0.39-0.73)* |
| -nurse | 1.17 (1.03-1.32)* | 1.16 (0.95-1.42) | 1.41 (1.19-1.68)* | 1.29 (0.94-1.77) | 1.03 (0.85-1.26) | 1.10 (0.83-1.45) |
| -auxiliary nurse | 1.74 (1.48-2.03)* | 1.73 (1.35-2.22)* | 1.81 (1.45-2.26)* | 1.62 (1.09-2.41)* | 1.72 (1.33-2.23)* | 1.79 (1.26-2.56)* |
| -other profession involved in patient care | 0.78 (0.66-0.93)* | 0.65 (0.48-0.88)* | 0.71 (0.55-0.91)* | 0.60 (0.37-0.98)* | 0.88 (0.67-1.15) | 0.70 (0.47-1.06) |
| -other profession not involved in patient care | Ref. | Ref. | Ref. | Ref. | Ref. | Ref. |
| Workplace setting |  |  |  |  |  |  |
| -hospital | 1.10 (0.95-1.28) | 1.08 (0.85-1.38) | 1.06 (0.86-1.30) | 1.07 (0.71-1.61) | 1.15 (0.91-1.47) | 1.14 (0.81-1.60) |
| -primary Care | 1.14 (0.95-1.36) | 1.14 (0.82-1.57) | 1.22 (0.94-1.58) | 1.24 (0.73-2.10) | 1.08 (0.82-1.44) | 1.11 (0.71-1.74) |
| -other | Ref. | Ref. | Ref. | Ref. | Ref. | Ref. |
| Frontline work during COVID-19 | 1.82 (1.66-1.99)* | 2.00 (1.72-2.32)* | 1.97 (1.73-2.23)* | 2.44 (1.89-3.14)* | 1.77 (1.54-2.04)* | 1.86 (1.53-2.27)* |
|  | Any current Mental Disorder (n=4,118) | Any current disabling Mental disorder (n=1,278) | Any current Mental Disorder (n=1,818) | Any current disabling Mental disorder (n=485) | Any current Mental Disorder (n=2,300) | Any current disabling mental disorder (n=793) |
|  | OR (95%CI) | OR (95%CI) | OR (95%CI) | OR (95%CI) | OR (95%CI) | OR (95%CI) |
| Frequency of direct exposure to COVID-19 patients |  |  |  |  |  |  |
| -all of the time | 3.30 (2.78-3.91)* | 3.88 (2.85-5.28)* | 4.37 (3.37-5.68)* | 5.78 (3.37-9.94)* | 3.11 (2.41-4.01)* | 3.68 (2.47-5.47)* |
| -most of the time | 2.53 (2.14-3.01)* | 3.27 (2.39-4.47)* | 3.04 (2.32-3.97)* | 4.35 (2.48-7.64)* | 2.42 (1.89-3.10)* | 3.04 (2.04-4.54)* |
| -some of the time | 2.01 (1.71-2.37)* | 2.24 (1.63-3.07)* | 2.49 (1.93-3.21)* | 2.70 (1.54-4.74)* | 1.88 (1.48-2.38)* | 2.21 (1.49-3.29)* |
| -a little of the time | 1.31 (1.09-1.58)* | 1.57 (1.13-2.20)* | 1.44 (1.08-1.91)* | 1.74 (0.96-3.16) | 1.42 (1.08-1.86)* | 1.73 (1.11-2.70)* |
| -none of the time | Ref. | Ref. | Ref. | Ref. | Ref. | Ref. |
| COVID-19 infection history |  |  |  |  |  |  |
| -having been hospitalized for COVID-19 | 1.41 (0.96-2.06) | 2.05 (1.23-3.41)* | 1.05 (0.59-1.85) | 1.83 (0.82-4.09) | 1.65 (0.90-3.02) | 2.18 (1.04-4.57)* |
| -positive COVID-19 test or medical COVID-19 diagnosis<sup>b</sup> | 1.07 (0.95-1.20) | 1.12 (0.93-1.34) | 1.03 (0.88-1.21) | 1.10 (0.82-1.48) | 1.11 (0.92-1.34) | 1.14 (0.88-1.47) |
| -none of the above | Ref. | Ref. | Ref. | Ref. | Ref. | Ref. |
| Having been isolated or quarantined because of COVID-19 | 1.35 (1.22-1.49)* | 1.52 (1.31-1.77)* | 1.27 (1.10-1.45)* | 1.52 (1.20-1.92)* | 1.41 (1.20-1.65)* | 1.56 (1.25-1.94)* |
| Close ones infected with COVID-19 |  |  |  |  |  |  |
| -partner, children, or parents | 1.25 (1.08-1.44)* | 1.19 (0.95-1.48) | 1.51 (1.25-1.84)* | 1.41 (1.01-1.96)* | 0.99 (0.79-1.25) | 0.97 (0.70-1.34) |
| -other family, friends or others<sup>c</sup> | 1.09 (0.98-1.20) | 0.98 (0.83-1.16) | 1.24 (1.08-1.42)* | 1.03 (0.78-1.35) | 0.95 (0.81-1.11) | 0.90 (0.71-1.14) |
| -none of the above | Ref. | Ref. | Ref. | Ref. | Ref. | Ref. |
| Lifetime mood disorder before onset COVID-19 outbreak | 3.18 (2.75-3.68)* | 4.88 (4.02-5.94)* | n.a. | n.a. | 1.61 (1.38-1.89)* | 2.41 (1.95-2.98)* |
| Lifetime anxiety disorder before onset COVID-19 outbreak | 3.15 (2.87-3.45)* | 3.72 (3.19-4.34)* | n.a. | n.a. | 1.52 (1.26-1.84)* | 1.44 (1.08-1.92)* |
| Lifetime substance use disorder before onset COVID-19 outbreak | 11.55 (6.18-21.56)* | 12.69 (6.17-26.11)* | n.a. | n.a. | 6.02 (3.21-11.28)* | 6.20 (2.91-13.19)* |
| Other lifetime mental disorder before onset COVID-19 outbreak | 1.62 (1.26-2.09)* | 2.32 (1.64-3.30)* | n.a. | n.a. | 0.79 (0.61-1.03) | 1.04 (0.72-1.50) |
| Any lifetime mental disorder before onset COVID-19 outbreak | 3.21 (2.93-3.51)* | 3.97 (3.40-4.63)* | n.a. | n.a. | n.a. | n.a. |
| Having two or more lifetime mental disorders (vs zero or exactly one) | 4.72 (3.95-5.65)* | 7.34 (5.85-9.21)* | n.a. | n.a. | 2.52 (2.09-3.05)* | 3.79 (2.98-4.84)* |

Note: OR, odds ratio; 95%CI, 95% confidence interval., n.a. not applicable
\* Statistically significant ( $\alpha=0.05$ ).
a All analyses adjust for time of survey (weeks), hospital membership and all predictors shown in the rows.
b the category "positive COVID-19 test or medical COVID-19 diagnosis" excludes those having been hospitalized for COVID-19.
c the category "other family, friends or others" excludes having a partner, children, or parents infected with COVID-19

**Table** 3 presents multivariable analyses of the associations described above, adjusting by all individual characteristics, COVID-19 exposure factors, and healthcare center and week of interview. Being female, and between ages 18-29 and being 30-49 were significantly associated with *any* and with *any disabling* current mental disorder. Being a physician and a nurse was consistently associated with significantly lower odds of current mental disorders, while being an auxiliary nurse with previous mental disorders showed high (but not significant) ORs of current disabling mental disorders. Being a frontline healthcare worker was a very important risk factor of any current and any disabling disorder, as it was also having been in quarantine or isolated. The factors most strongly associated with current disabling mental disorders were previous substance use disorders, anxiety disorder and depression disorders. Having more than one previous disorders was no longer statistically significant in the multivariate analysis.

**Table 3.** Multivariable associations between individual characteristics, COVID-19 exposure, and prior lifetime (LT) mental disorders with current mental disorders and current disabling mental disorders. Spanish healthcare workers, MINDCOVID study (N=9,138).

|  | ALL<br>(n=9,138) |  | No Prior LT Mental Disorders<br>(New Onset) (n=5,367) |  | Prior LT Mental Disorder<br>(Persistence/Relapse) (n=3,771) |  |
| --- | --- | --- | --- | --- | --- | --- |
|  | Any current Mental Disorder (n=4,118) | Any current disabling Mental disorder (n=1,278) | Any current Mental Disorder (n=1,818) | Any current disabling Mental disorder (n=485) | Any current Mental Disorder (n=2,300) | Any current disabling Mental disorder (n=793) |
|  | aOR (95%CI) | aOR (95%CI) | aOR (95%CI) | aOR (95%CI) | aOR (95%CI) | aOR (95%CI) |
| Gender - female (vs male) | 1.45 (1.29-1.63)* | 1.58 (1.27-1.96)* | 1.54 (1.31-1.81)* | 1.77 (1.27-2.46)* | 1.36 (1.13-1.63)* | 1.50 (1.12-2.01)* |
| Age |  |  |  |  |  |  |
| -18-29 years | 1.53 (1.28-1.82)* | 1.36 (1.02-1.82)* | 1.82 (1.43-2.32)* | 1.67 (1.07-2.61)* | 1.23 (0.95-1.59) | 1.17 (0.80-1.71) |
| -30-49 year | 1.46 (1.30-1.64)* | 1.34 (1.09-1.64)* | 1.49 (1.27-1.74)* | 1.39 (1.04-1.87)* | 1.42 (1.19-1.70)* | 1.30 (0.98-1.74) |
| -50 years or more | Ref. | Ref. | Ref. | Ref. | Ref. | Ref. |
| Marital status - married (vs single, divorced or legally separated, or widowed) | 1.05 (0.95-1.17) | 1.02 (0.85-1.23) | 1.09 (0.94-1.26) | 1.09 (0.84-1.42) | 1.01 (0.86-1.19) | 0.98 (0.77-1.26) |
| Having children in care |  |  |  |  |  |  |
| -younger (<12 ys) children in care | 0.97 (0.85-1.11) | 1.01 (0.81-1.26) | 1.00 (0.84-1.20) | 1.03 (0.74-1.42) | 0.93 (0.76-1.13) | 0.99 (0.73-1.33) |
| -children in care, but >12ys | 0.96 (0.84-1.11) | 0.89 (0.70-1.13) | 0.95 (0.78-1.14) | 0.82 (0.57-1.16) | 0.99 (0.80-1.24) | 0.95 (0.68-1.32) |
| -no children in care | Ref. | Ref. | Ref. | Ref. | Ref. | Ref. |
| Profession |  |  |  |  |  |  |
| -physician | 0.45 (0.39-0.53)* | 0.34 (0.26-0.44)* | 0.45 (0.37-0.55)* | 0.30 (0.20-0.45)* | 0.45 (0.36-0.56)* | 0.36 (0.25-0.52)* |
| -nurse | 0.77 (0.67-0.90)* | 0.73 (0.57-0.92)* | 0.82 (0.67-0.99)* | 0.69 (0.48-0.98)* | 0.70 (0.56-0.87)* | 0.73 (0.53-1.01) |
| -auxiliary nurse | 1.12 (0.94-1.34) | 1.07 (0.80-1.42) | 1.11 (0.87-1.41) | 0.92 (0.60-1.40) | 1.13 (0.85-1.50) | 1.18 (0.79-1.78) |
| -other profession involved in patient care | 0.67 (0.55-0.80)* | 0.54 (0.39-0.75)* | 0.59 (0.45-0.77)* | 0.50 (0.30-0.82)* | 0.75 (0.56-1.00) | 0.57 (0.36-0.92)* |
| -other profession not involved in patient care | Ref. | Ref. | Ref. | Ref. | Ref. | Ref. |
| Frequency of direct exposure to COVID-19 patients |  |  |  |  |  |  |
| -all of the time | 3.98 (3.27-4.85)* | 5.19 (3.61-7.46)* | 4.40 (3.31-5.85)* | 6.62 (3.70-11.85)* | 3.53 (2.66-4.68)* | 4.27 (2.70-6.76)* |
| -most of the time | 3.10 (2.54-3.76)* | 4.53 (3.17-6.48)* | 3.15 (2.36-4.20)* | 5.00 (2.77-9.01)* | 3.08 (2.34-4.06)* | 4.26 (2.70-6.71)* |
| -some of the time | 2.50 (2.08-3.01)* | 2.96 (2.07-4.23)* | 2.74 (2.09-3.60)* | 3.25 (1.82-5.82)* | 2.24 (1.73-2.90)* | 2.71 (1.74-4.20)* |
| -a little of the time | 1.55 (1.27-1.90)* | 1.93 (1.33-2.80)* | 1.60 (1.19-2.14)* | 2.06 (1.13-3.78)* | 1.53 (1.15-2.03)* | 1.81 (1.12-2.92)* |
| -none of the time | Ref. | Ref. | Ref. | Ref. | Ref. | Ref. |
| COVID-19 infection history |  |  |  |  |  |  |
| -having been hospitalized for COVID-19 | 1.06 (0.69-1.64) | 1.42 (0.76-2.65) | 0.92 (0.50-1.71) | 1.48 (0.60-3.65) | 1.23 (0.64-2.39) | 1.38 (0.57-3.34) |
| -positive COVID-19 test or medical COVID-19 diagnosis<sup>b</sup> | 0.82 (0.70-0.95)* | 0.76 (0.60-0.96)* | 0.77 (0.63-0.93)* | 0.69 (0.48-1.01) | 0.88 (0.70-1.11) | 0.83 (0.60-1.14) |
| -none of the above | Ref. | Ref. | Ref. | Ref. | Ref. | Ref. |
|  | Any current Mental Disorder (n=4,118) | Any current disabling Mental disorder (n=1,278) | Any current Mental Disorder (n=1,818) | Any current disabling Mental disorder (n=485) | Any current Mental Disorder (n=2,300) | Any current disabling Mental disorder (n=793) |
|  | aOR (95%CI) | aOR (95%CI) | aOR (95%CI) | aOR (95%CI) | aOR (95%CI) | aOR (95%CI) |
| Having been isolated or quarantined because of COVID-19 | 1.36 (1.20-1.54)* | 1.60 (1.31-1.95)* | 1.34 (1.13-1.59)* | 1.64 (1.20-2.25)* | 1.41 (1.16-1.71)* | 1.60 (1.21-2.11)* |
| Lifetime mood disorder before onset COVID-19 outbreak | 2.53 (2.04-3.15)* | 3.23 (2.27-4.60)* | n.a. | n.a. | 2.91 (1.29-6.57)* | 4.46 (1.65-12.03)* |
| Lifetime anxiety disorder before onset COVID-19 outbreak | 2.82 (2.53-3.13)* | 3.03 (2.53-3.62)* | n.a. | n.a. | 3.36 (1.49-7.58)* | 4.30 (1.61-11.43)* |
| Lifetime substance use disorder before onset COVID-19 outbreak | 8.25 (4.22-16.11)* | 5.74 (2.53-13.03)* | n.a. | n.a. | 9.17 (3.67-22.92)* | 7.23 (2.60-20.08)* |
| Other lifetime mental disorder before onset COVID-19 outbreak | 1.53 (1.15-2.05)* | 2.06 (1.35-3.13)* | n.a. | n.a. | 1.77 (0.80-3.89) | 2.69 (1.07-6.76)* |
| Having two or more lifetime mental disorders (vs zero or exactly one) | 0.93 (0.69-1.24) | 1.17 (0.76-1.79) | n.a. | n.a. | 0.78 (0.33-1.83) | 0.83 (0.29-2.36) |
| AUC | 0.73 | 0.77 | 0.68 | 0.72 | 0.69 | 0.73 |

aOR, adjusted Odd Ratio; CI, 95% Confidence Interval; ref, reference category; AUC, Area under the curve. SE, Standard Error, n.a.not applicable
\* Statistically significant ( $\alpha=0.05$ ).
a Each column represents a separate regression model, each time adjusting for time of survey (weeks), hospital membership, and all
b the category "positive COVID-19 test or medical COVID-19 diagnosis" excludes those having been hospitalized for COVID-19.

## DISCUSSION

Our results document a high prevalence of current mental disorders, with almost half of respondents screening positive on at least one of the five well-established screeners for common mental disorders. Most important, 1 in 7 met criteria for a current disabling mental disorder. To the best of our knowledge, this is the first study to consider both symptom screening and disability as indicator of adverse mental health during the COVID-19 pandemic. Such a combination is potentially more valid and useful for services planning purposes, than descriptive information on psychological symptoms^34,35^. We also found that prevalence of adverse mental health was significantly more frequent among healthcare workers with prior mental disorders. Finally, we found that being a female, having a high frequency of exposure to COVID-19 patients, and having quarantined or isolated are risk factors for both any current disorder and any disabling disorder.

### Comparison with other studies

The prevalence estimates found in our study of MDD (28.1%) and GAD (22.5%) was well within the range of meta-analytic reports of healthcare workers studied in predominantly Asian healthcare settings^6,8-9^. Our estimated prevalence of PTSD (22.2%) was also remarkably similar to that reported among healthcare workers in a meta-analysis (20.7%)^5^. Substance use disorder was present in 6.2% of our sample. Only a few studies have reported empirical estimates of this disorder during the COVID-19 pandemic and we were unable to find any specific data among healthcare workers. Our results suggest that this disorder has a considerably lower prevalence than found in the general adult populations of the US^36^ and France^37^.

To the best of our knowledge, no previous report has presented data on the prevalence of any mental disorder and any disabling mental disorder among healthcare workers during the COVID-19 pandemic. The prevalence in our study (i.e., 45.7% of the responding healthcare workers meet criteria for any of the five assessed disorders) is somewhat higher than the 40.9% of ≥ 1 adverse mental or behavioral health symptom in the adult US population^36^. More importantly, 1 in 7 presented a current disabling mental disorder, pointing to the high interference of adverse mental health on, professional, domestic, personal, and social activities. Our results suggest that there are large mental healthcare needs to meet among healthcare professionals. There is need to closely monitor the extent to which these needs are adequately met.

An important finding of our study is the strong association of prior lifetime disorders with any current disabling mental disorder (with odds ratios ranging from 1.53 to 8.25). This result, which is consistent with our clinical experience during the first wave of the pandemic, strongly suggests that healthcare workers with such a history must be considered a group at especially high risk. Adequate mental health monitoring and support measures should be made accessible to this important group of healthcare workers.

### Strengths and limitations

Strengths of our study include the large number of institutions included and their spread over the most affected regions of Spain; the use of the institutional mailing lists as the sampling framework, which provided specific and reliable listing of healthcare workers; data representative of a large number of healthcare workers; and the higher validity of screening of symptoms with severe interference to identify disabling mental disorders. These strengths support the robustness and relevance of our results.

Nevertheless, the study has some limitations that deserve careful consideration. First, we had a low response rate. Despite important advantages of institutional email listings, these email accounts seem not to be checked by a large majority of employees and their utilization might differ by professional category. In fact, in our study we could assess the proportion of workers who read their first email invitation, which was less than 27%. In addition, invitations were limited to a maximum of 2 due to institutional requirements. However, in order to improve representativeness, we have carefully weighted the observed data as to exactly reproduce the gender, age and professional category distribution of healthcare personnel in each participating institution.

Second, the study was cross-sectional in nature and it cannot be used to infer any causal impact of the COVID-19 pandemic on the mental health of healthcare workers. Nevertheless, we used clear and relevant recall periods to make sure the symptoms were present after the pandemic and had started for most of the symptoms, a short period before the interview. Furthermore, we did collect information on mental disorders the respondents had suffered any time in their life before the COVID-19 pandemic.

Third, measures used to assess mental disorders in our study are based on self-reports and not on clinical diagnoses. Nevertheless, there is good evidence of acceptable sensitivity and specificity of the assessment for the current score cutoffs used here for current major depression disorder^15^, generalized anxiety disorder^16^ and post-traumatic stress disorders^19^. These measures are among the most frequently used in epidemiologic studies which allows comparability of results. For lifetime disorders we used a list of disorders which have been shown to have acceptable agreement with clinical evaluations^38^. The high prevalence of both lifetime and current mental disorders found in our study suggests that a part might include false positive cases; and some of the real cases may have a mild disorder. It is for this reason that we propose to consider disabling current mental disorders a better estimate the needs for mental healthcare in this population^34,35^. Healthcare workers with disabling current mental disorder in our study had much more frequent (between 2 and three times more) mental comorbidity, current suicidal ideation, poor perceived (data not presented, available upon request).

## Conclusions

Notwithstanding the limitations, our study shows a high prevalence of current mental disorders among Spanish healthcare workers during the first wave of the COVID-19 pandemic, with 1 in 7 presenting a disabling mental disorder. Prevalence of adverse mental health was significantly more frequent among healthcare workers reporting lifetime mental disorders before the pandemic, which identifies a group in need of current monitoring and adequate support, especially as the pandemic is entering in its second wave. Other healthcare workers that should be monitored include with a high frequency of exposure to COVID-19 patients, who had been infected or have been quarantined or isolated, as well as female workers, auxiliary nurses and nurses.

## Data Availability

The de-identified participant data as well as the study protocol and statistical analysis plan used for this study are available upon reasonable request from the corresponding author (JA) as long as the main objective of the data sharing request is replicating the analysis and findings as reported in this paper.

## ACKNOWLEDGMENTS

The authors would like to sincerely thank all healthcare workers that participated in the study in extremely busy times. They also thank very much Puri Barbas and Franco Amigo for the management of the project, and Carme Gasull for manuscript preparation and submission.

