## Supplemental Table 1 for "Mental Health Impact of the First Wave of COVID-19 Pandemic on Spanish Healthcare Workers: a Large Cross-sectional Survey"

**Supplementary Table 1.Current prevalence of mental disorder stratified by participating health center (N = 9,138).**

|  | | | **Current MDD (n = 2554)** | **Current GAD (n = 2007)** | **Current Panic Attacks (n = 2064)** | **Current PTSD (n = 1946)** | **Current Substance Use Disorder (n = 569)** | **Any current Mental Disorder (n = 4118)** | **Any current disa Mental Disorder (n=1278)** | **Response**  **rate^a^** | **Participation rate^b^** | **Completion rate^c^** |
| --- | --- | --- | --- | --- | --- | --- | --- | --- | --- | --- | --- | --- |
|  | **n** | **%** | **% (SE)** | **% (SE)** | **% (SE)** | **% (SE)** | **% (SE)** | **% (SE)** | **% (SE)** | **%** | **%** | **%** |
| Total |  |  | 28.1 (0.5)* | 22.5 (0.4)* | 24.0 (0.5)* | 22.2 (0.4)* | 6.2 (0.3)* | 45.7 (0.5)* | 14.5 (0.5)* | 12.50 | 89.65 | 80.60 |
| H. Center 1 | 452 | 5.8 (0.2)* | 23.1 (1.8)* | 20.0 (1.8)* | 21.7 (1.8)* | 22.2 (1.8)* | 5.7 (1.1) | 43.7 (2.2)* | 14.5 (1.8)* | 7.60 | 87.11 | 75.9 |
| H. Center 2 | 614 | 8.4 (0.3) | 42.5 (1.8) | 33.6 (1.7) | 34.6 (1.8) | 30.7 (1.7) | 6.3 (0.9) | 60.1 (1.8) | 18.2 (1.9) | 7.00 | 88.16 | 81.19 |
| H. Center 3 | 238 | 2.0 (0.1) | 21.1 (3.1) | 17.5 (2.9) | 17.6 (2.9) | 18.3 (2.9) | 3.7 (1.4) | 35.2 (3.6) | 8.7 (2.4) | 16.70 | 86.07 | 77.62 |
| H. Center 4 | 870 | 7.1 (0.3) | 33.6 (1.9) | 25.5 (1.7) | 26.9 (1.8) | 27.2 (1.8) | 5.0 (0.9) | 51.2 (2.0) | 16.3 (1.6) | 11.80 | 88.47 | 78.92 |
| H. Center 5 | 1015 | 4.3 (0.2) | 29.4 (2.3) | 23.0 (2.1) | 26.4 (2.2) | 20.8 (2.0) | 6.3 (1.2) | 46.5 (2.5) | 13.8 (1.8) | 22.50 | 88.43 | 81.91 |
| H. Center 6 | 805 | 18.7 (0.4) | 24.5 (1.0) | 21.1 (1.0) | 24.5 (1.1) | 18.8 (1.0) | 5.3 (0.6) | 42.6 (1.2) | 12.8 (1.2) | 4.10 | 89.49 | 83.86 |
| H. Center 7 | 85 | 3.8 (0.2) | 13.8 (1.9) | 11.6 (1.7) | 16.2 (2.1) | 13.4 (1.8) | 4.9 (1.2) | 27.3 (2.5) | 9.4 (2.1) | 2.20 | 85.32 | 83.87 |
| H. Center 8 | 66 | 0.1 (0.0) | 27.3 (13.7) | 17.8 (11.8) | 11.4 (9.8) | 16.1 (11.3) | 6.4 (7.5) | 35.2 (14.7) | 15.2 (11.2) | 53.50 | 97.06 | 93.94 |
| H. Center 9 | 618 | 6.3 (0.3) | 23.1 (1.8) | 17.7 (1.6) | 20.5 (1.7) | 19.4 (1.7) | 6.4 (1.0) | 42.2 (2.1) | 11.4 (1.5) | 9.20 | 91.87 | 83.19 |
| H. Center 10 | 1695 | 15.0 (0.4) | 32.0 (1.3) | 25.1 (1.2) | 22.1 (1.1) | 24.6 (1.2) | 6.2 (0.7) | 47.5 (1.4) | 16.2 (1.2) | 15.80 | 92.3 | 82.61 |
| H. Center 11 | 224 | 1.1 (0.1) | 18.3 (3.9) | 15.1 (3.6) | 13.0 (3.4) | 12.0 (3.2) | 2.4 (1.5) | 29.4 (4.6) | 7.2 (3.0) | 19.40 | 89.9 | 81.4 |
| H. Center 12 | 300 | 3.9 (0.2) | 32.3 (2.5) | 25.5 (2.3) | 30.5 (2.5) | 21.0 (2.2) | 7.2 (1.4) | 52.3 (2.7) | 16.9 (2.4) | 7.40 | 94.07 | 83.78 |
| H. Center 13 | 97 | 4.2 (0.2) | 29.1 (2.3) | 20.1 (2.1) | 18.3 (2.0) | 25.9 (2.3) | 9.4 (1.5) | 45.7 (2.6) | 12.8 (3.1) | 2.30 | 92.86 | 79.81 |
| H. Center 14 | 608 | 5.5 (0.2) | 20.8 (1.8) | 15.1 (1.6) | 17.6 (1.7) | 18.5 (1.7) | 7.2 (1.2) | 38.5 (2.2) | 12.2 (1.7) | 11.00 | 88.4 | 73.34 |
| H. Center 15 | 343 | 2.6 (0.2) | 34.4 (3.1) | 30.6 (3.0) | 29.5 (3.0) | 32.3 (3.0) | 7.5 (1.7) | 58.5 (3.2) | 21.3 (2.9) | 12.90 | 88.41 | 74.36 |
| H. Center 16 | 616 | 5.1 (0.2) | 21.9 (1.9) | 15.4 (1.7) | 21.5 (1.9) | 15.6 (1.7) | 9.1 (1.3) | 40.7 (2.3) | 10.8 (1.7) | 11.40 | 91.42 | 80.28 |
| H. Center 17 | 119 | 0.5 (0.1) | 27.8 (6.8) | 20.5 (6.1) | 14.1 (5.4) | 23.3 (6.4) | 6.2 (3.7) | 42.0 (7.5) | 8.9 (4.7) | 23.70 | 90.48 | 88.16 |
| H. Center 18 | 373 | 5.7 (0.2) | 30.7 (2.0) | 28.0 (2.0) | 28.0 (2.0) | 25.1 (1.9) | 5.3 (1.0) | 49.6 (2.2) | 20.8 (2.1) | 6.30 | 84.97 | 79.25 |
| p-value |  | <0.001* | <0.001* | <0.001* | <0.001* | <0.001* | 0.08 | <0.001* | <0.001* |  | <0.001* | <0.001* |

* Statistically significant (α=0.05).
a Response rate is calculated as the number of participants that completed survey section A (sociodemographic) and B (COVID-19 infection status) divided by the estimated total number of eligible hospital workers. The total number of eligible hospital workers was estimated using the size of the hospitals' email distribution lists, or by the total number of hospital workers employed as provided by the healthcare centers. The total response rate is adjusted by achieved sample size; the unadjusted total response rate is 9.2%.
b The participation rate is calculated as the number of hospital workers that agreed to participate in the survey divided by the total number of hospital workers that responded to the informed consent (i.e., the first page of the web-based survey).
c The completion rate is calculated as the number of participants that completed the survey divided by the participants that agreed to participate in the survey.
